# Glucose-lowering effects of physical activity in type 1 diabetes: A causal modelling and matched-pair analysis approach

**DOI:** 10.1101/2025.05.29.25328403

**Authors:** Catherine L. Russon, John S Pemberton, Richard M. Pulsford, Brad S. Metcalf, Emma Cockcroft, Michael J Allen, Anne M Frohock, Rob C. Andrews

## Abstract

**OBJECTIVE:** To evaluate the acute glucose-lowering effect of bouts of physical activity (PA) for hyperglycemia in individuals with type 1 diabetes (T1D), using a within-subject matched-pairs causal design to approximate the control condition of no activity.

**RESEARCH DESIGN AND METHODS:** Data comprised 1546 PA bouts of 10-30 min from 482 participants in the T1DEXI and T1DEXIP cohorts where glucose was >180 mg/dL (10 mmol/L). Each PA bout was matched [starting glucose, glucose rate of change, insulin on board (IOB), and glucose variability (CV) to a matched non-PA period within the same individual using a weighted k-nearest neighbors algorithm (SMD <0.01). Primary outcome: Change in glucose from PA onset to 20 minutes post-activity. Secondary outcomes: Predictors of glucose response and rate of hypoglycemia incidence.

**RESULTS:** PA [median 23 minutes: IQR (20, 30)] led to a mean glucose change of -40 mg/dL (-2.2 mmol/L, p<0.001), compared to -5 mg/dL (0.3 mmol/L, p<0.001) during matched non-PA periods (mean difference: -35 mg/dL[1.9 mmol/L], p<0.0001). No significant differences by age, activity type, or intensity were observed. The strongest predictors of PA-induced glucose change were (in order) glucose rate of change, starting glucose, CV, duration, and IOB. A heatmap using starting glucose and glucose rate of change was developed to guide real-time decision-making. PA-induced hypoglycemia risk was very low (<2%).

**CONCLUSIONS:** Using PA to lower high glucose levels is an effective and safe strategy, and when guided by CGM, it can become a personalized tool for T1D education.

**ARTICLE HIGHLIGHTS:** *Why did we undertake this study?:* To test whether short bouts of physical activity can safely and effectively lower high glucose levels in people with type 1 diabetes.

*What specific question(s) did we want to answer?:* Does 10–30 minutes of physical activity, started when glucose is high, reduce glucose more than doing nothing, and what factors affect this response?

*What did we find?:* Across 1,546 activity bouts, 23 minutes of exercise lowered glucose by 40 mg/dL (2.2 mmol/L)—eight times more than rest—with <2% hypoglycemia risk.

*What are the implications of our findings?:* Short bouts of activity quickly and safely lower high glucose, with drop size best predicted by starting level and trend—making it easy to teach and implement using a simple heatmap.

**Twitter Summary:** Short bouts of physical activity (10–30 min) lower glucose by 40 mg/dL (2.2 mmol/L) during hyperglycemia in people with type 1 diabetes, compared to 5 mg/dL (0.3 mmol/L) with no activity. Top predictors: starting glucose, rate of change, variability, duration, insulin on board. Hypoglycemia risk was very low (<2%). CGM-guided activity is a safe, effective, and personalisable glucose-lowering tool. #T1D #CGM #Exercise

**Graphical Abstract:** 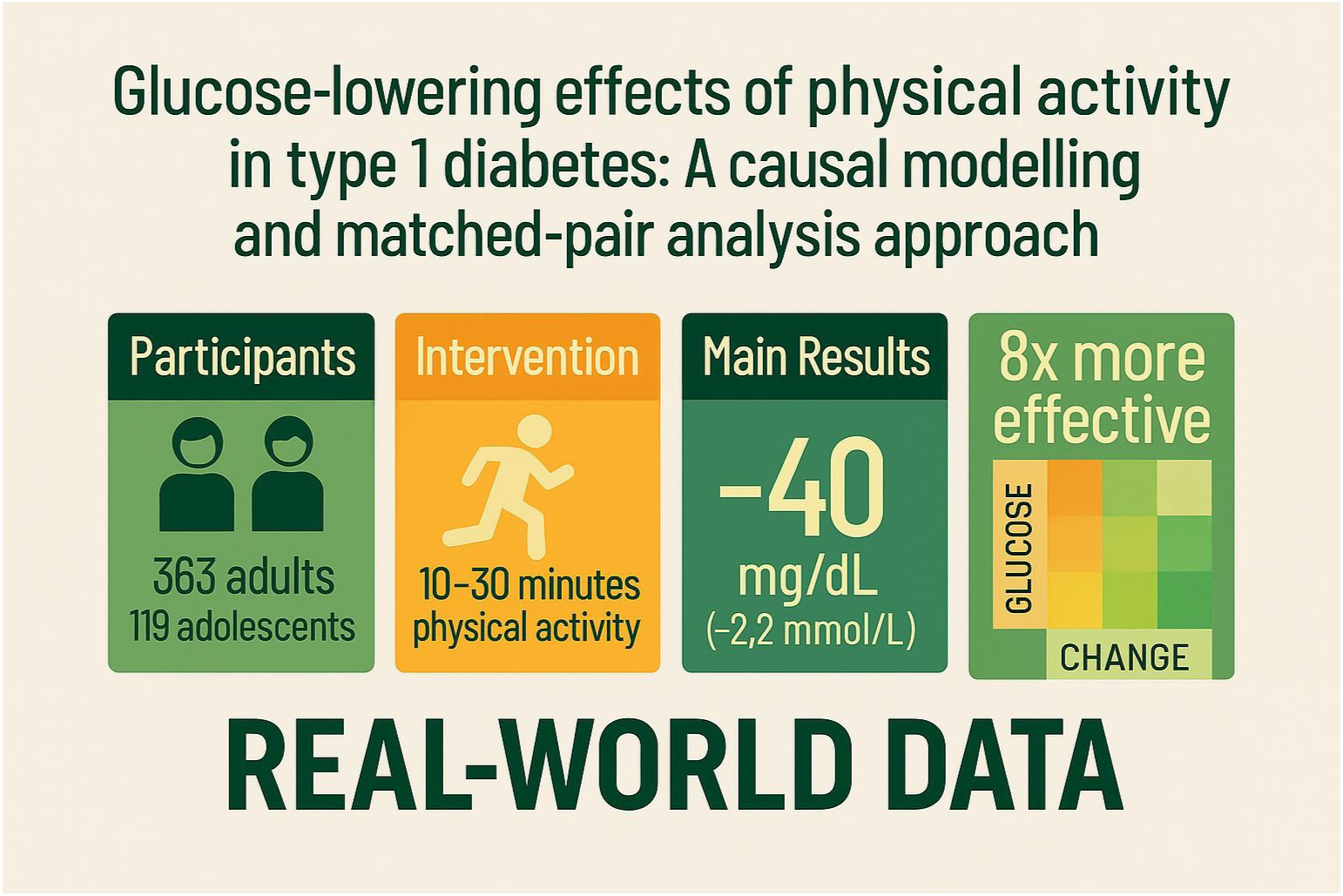

---

Achieving optimal glycemic control in type 1 diabetes (T1D) remains a significant challenge. Despite widespread use of continuous glucose monitoring (CGM) and hybrid closed-loop (HCL) systems, most users fail to reach the recommended target of ≥70% time in range (TIR; 70–180 mg/dL, 3.9-10 mmol/L) (1–3). Hyperglycemia is usually corrected with rapid-acting insulin, but its delayed onset and prolonged action often result in slow and unpredictable glucose responses (4). Bolus calculators often underestimate insulin action time, driving further insulin delivery and increasing the risk of insulin stacking and subsequent hypoglycemia (5). Ongoing exposure to hyperglycemia increases the risk of serious long-term complications, including microvascular and macrovascular damage (6,7). These challenges highlight the urgent need for complementary strategies to improve glucose control and reduce the burden of disease.

The longer-term health benefits of physical activity (PA) in T1D are well established, and PA is recommended as a cornerstone of diabetes management. Alongside its long-term influence on health outcomes, PA can serve as a non-pharmacologic strategy for hyperglycemia management, lowering glucose through insulin-independent mechanisms and enhancing insulin sensitivity (8,9). Experimental data demonstrate that PA in controlled environments can reduce glucose by 36–144 mg/dL (2-8 mmol/L) within two hours of a meal (10). However, these small experimental studies often involve highly selected samples and standardized exercise protocols, limiting external validity and providing little insight into the effects of heterogeneous exercise behaviors in uncontrolled, free-living environments. Observational real-world studies provide some evidence of efficacy in lowering glucose levels above 180 mg/dL (10 mmol/L) (11), but the absence of counterfactual comparators in these studies limits causal inference and undermines the application of PA as an acute glucose-lowering intervention.

Furthermore, although the benefits of short bouts of PA are beginning to be established (12), little work has translated these insights into practical tools to support implementation in daily life.

Causal modelling offers a reliable way to estimate the effects of different factors in observational data by mimicking the conditions of a randomized controlled trial (13). In diabetes research, causal modelling has been used to explore cause-and-effect relationships while reducing the impact of confounding factors and reverse causation (14).

To date, no studies have applied causal inference methods to evaluate the potential of PA to acutely lower elevated glucose levels, using continuous glucose monitoring (CGM) data collected under free-living conditions. Applying causal modeling to these events, while accounting for both individual and event-specific factors, can generate robust evidence regarding the impact of everyday behaviors in real-world settings, supporting the development of new evidence-based interventions.

This study employed a novel within-subject matched-pairs approach applied to the T1DEXI (15) and T1DEXIP (16) datasets, intending to provide the first causal evidence for the glucose-lowering effect of free-living PA in individuals with T1D. By comparing each bout of PA to a matched non-PA period within the same individual (matched on key person-level and event-level variables), this design helps estimate what would have happened without the activity, thereby reducing the influence of other factors that could affect the results.

The primary aim was to compare the change in glucose concentration from activity onset to 20 minutes post-activity for 10–30 minute bouts of PA initiated at glucose levels >180 mg/dL (10 mmol/L). The secondary aims were to identify moderators of glucose responses between PA and matched non-PA periods and to assess the incidence of hypoglycemia for the same period compared with matched non-PA periods under free-living conditions. Finally, the study explored whether these findings could be used to develop a simple, user-friendly tool to help individuals with T1D understand the likely glucose-lowering effect of physical activity at different starting glucose levels and rates of glucose change.

## Research Design and Methods

An overview of the methods used in this study is available in **Fig. 1**. This study is reported in accordance with the **STROBE guidelines** for observational research.

**Figure 1.**
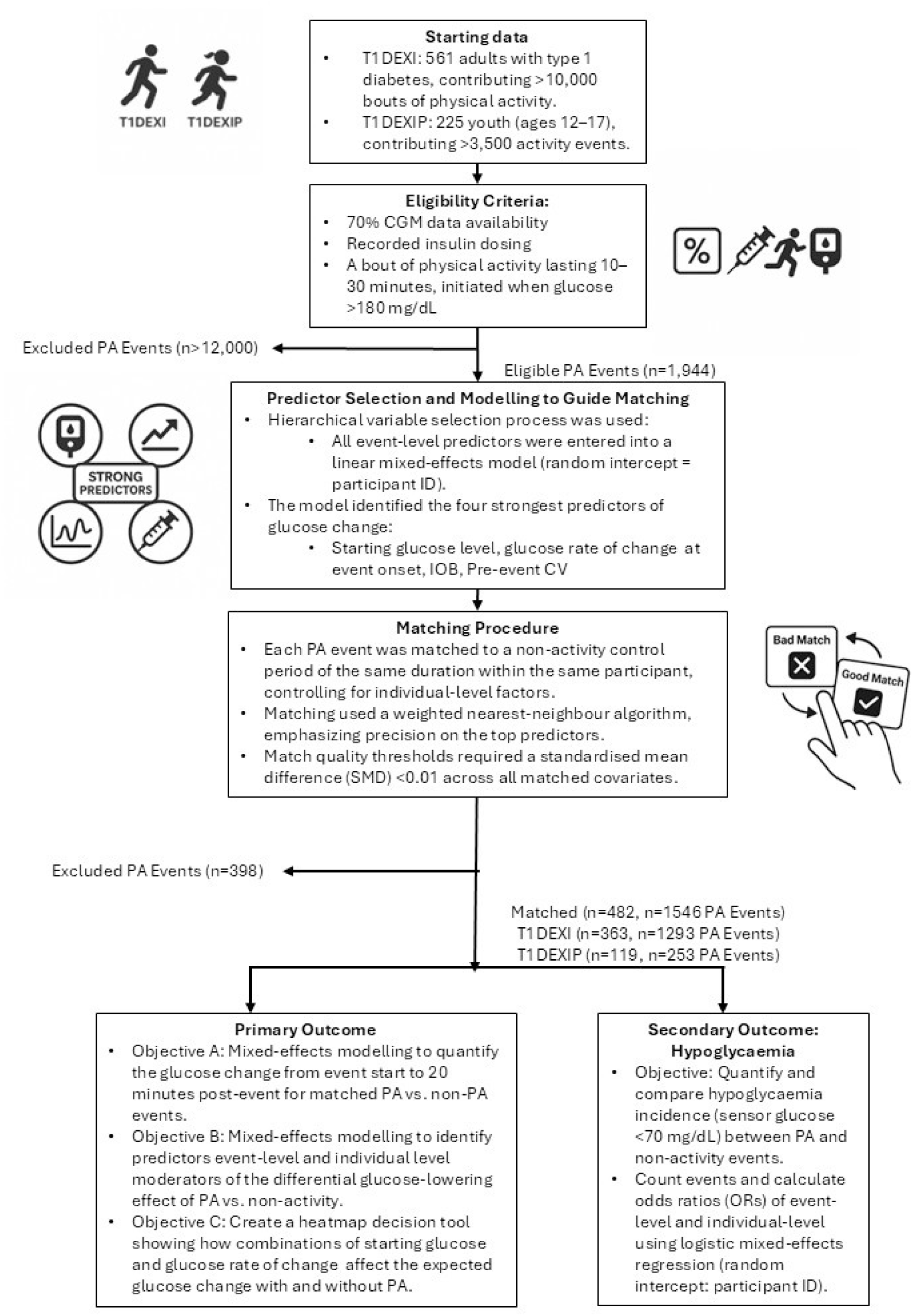
Overview of the Analysis Plan Using Matched-Pairs Design. This flowchart outlines the analytical workflow for evaluating the glucose-lowering effect of physical activity (PA) in type 1 diabetes (T1D) using a within-subject matched-pairs design. PA refers to physical activity, and T1D stands for type 1 diabetes. T1DEXI and T1DEXIP refer to the Type 1 Diabetes Exercise Initiative adult and pediatric cohorts, respectively. IOB represents insulin on board at the start of the event. CV is the coefficient of variation of glucose in the hour prior to activity, and SMD refers to the standardised mean difference used to assess the quality of matching between PA and non-PA.

### Study Population

Participants were drawn from two real-world cohorts of individuals with Type 1 diabetes: T1DEXI (561 adults, >10,000 PA bouts) and T1DEXIP (225 adolescents aged 12–17 years, >3,500 PA bouts) (12–14). All participants were using continuous glucose monitoring and insulin therapy in free-living conditions. Eligibility for this analysis required 70% CGM data availability, insulin dosing data, with at least one qualifying bout of PA lasting 10–30 minutes and initiated at glucose >180 mg/dL (10 mmol/L). Only PA bouts with sufficiently matched non-PA periods were included to allow for robust within-subject comparisons. Full cohort characteristics and data collection methods are detailed in the original publications (15,16).

### Outcomes

The primary outcome was the change in sensor glucose from the start of PA to 20 minutes post-activity, accounting for CGM lag (17), comparing 10–30 minute PA bouts to matched non-PA periods. All bouts began at a glucose level >180 mg/dL (10 mmol/L) under free-living conditions. The secondary outcome was assessing hypoglycemia incidence (<70 mg/dL, <3.9 mmol/L) during or within 20 minutes after PA, compared to matched control periods.

### Causal Matched-Pairs Design

Rather than relying on propensity scores, which have well-documented limitations for causal inference in matched designs (18), we adopted an outcome-oriented approach. Specifically, we prioritized matching on covariates that most strongly influenced glucose change following PA. This is conceptually aligned with prognostic score matching, which has been shown to reduce bias more effectively when the goal is to estimate causal effects on an outcome (19,20).

To identify covariates for matching, we conducted a variable selection process. Variables significantly associated with glucose change in univariate Pearson correlations were entered into a multivariable linear mixed-effects model with a random intercept for participant ID. Variables retained in this model were used for matching.

Each PA bout was matched to a non-PA control period of equal duration, drawn from the same participant’s CGM data. This within-subject design inherently controlled for baseline characteristics such as age, sex, insulin modality, and HbA1c.

Matching was performed using a weighted k-nearest neighbors (kNN) algorithm (20). Variable weights were derived from the coefficients of the multivariable model, prioritizing tighter matches on more influential variables, while allowing greater flexibility for less predictive covariates. A Euclidean distance threshold was set to ensure poor-quality matches were excluded.

To minimise confounding, overnight periods and time windows within 1 hour before or 4 hours after PA bouts were excluded. A buffer of 40 minutes on either side was also applied to prevent overlap between matched periods.

Final match quality was assessed using standardised mean differences (SMDs), with values below 0.1 considered indicative of acceptable covariate balance (21).

### Statistical Analysis

Descriptive statistics were reported as the mean (± SD), median (IQR: 25th, 75th), or n (%), as appropriate. Glucose changes are reported as means with 95% confidence intervals (95% CI). Quadratic and cubic terms were tested alongside standard terms at every stage of analysis. Two-sided p-values <0.05 were considered statistically significant.

A three-level linear mixed-effects model was used to estimate the association between PA and glucose change. Each observation was nested within a matched PA bout/non-PA period (Level 2), which was nested within participant (Level 3). The model included fixed effects for PA condition and relevant covariates (see **Supplementary Data: S1-3**), as well as interaction terms to assess whether the PA effect on glucose changes varied by physiological state. A random slope for PA was included at the participant level to allow for individual variation in glycemic response, and a random intercept for matched bouts was included to account for pairing structure. This approach accounts for differences between individuals and bouts unexplained by other variables in the model.

Hypoglycemia risk was modelled using a logistic mixed-effects regression with fixed effects for covariates and a random intercept for participants to account for within-subject clustering. Odds ratios (ORs) were estimated to compare the likelihood of hypoglycemia between activity conditions.

All analyses were performed in Python (v3.9) and R (v4.3.1), using the Diametrics (22), lme4 (23), lmerTest, Pandas (24), Numpy, and statsmodels packages (25). All code is available at https://github.com/cafoala/glucoseLo-stats.

### Ethics Statement

This study involved secondary analysis of de-identified data from the T1DEXI and T1DEXIP studies, both of which obtained institutional review board approval and informed consent from all participants, as described in the original publications (15,16).

This secondary analysis was conducted in accordance with the principles of the Declaration of Helsinki.

## Results

### Participant Characteristics

A total of 482 participants were included in the analysis, comprising 363 from the T1DEXI cohort and 119 from T1DEXIP. The T1DEXI group had a median age of 32 years (IQR: 25, 45), compared to 13 years (12, 14) in the T1DEXIP group. In the T1DEXI cohort, 44% of participants were using HCL systems, 41% were on standard insulin pumps, and 15% were managing diabetes via MDI. For the T1DEXIP group, 56% were using HCL systems, with 29% on pumps and 15% using MDI. Full demographic details are available in **Table 1**.

**Table 1.** Participant Characteristics, Baseline glycemic Metrics, and Physical Activity Bout Features in the Overall Cohort and by Study Group (T1DEXI vs. T1DEXIP) Data are presented as median [interquartile range] or n (%), as appropriate. T1DEXI participants represent the adult cohort, while T1DEXIP participants reflect the adolescent/paediatric cohort. glycemic metrics are derived from continuous glucose monitoring (CGM) data, and include measures of average glucose, glycemic variability (SD, CV, MAGE), and time in range (TIR). Physical activity (PA) characteristics reflect matched activity and non-PA bout pairs, including starting glucose, insulin on board (IOB), bout duration, and contextual variables such as PA type, intensity, and time of day. CV = coefficient of variation.

|  | Overall | T1DEXI | T1DEXIP |  |  |  |
| --- | --- | --- | --- | --- | --- | --- |
| Demographic Characteristics |  |  |  |  |  |  |
| Number of participants | 482 | 363 | 119 |  |  |  |
| Age | 26 [18, 39] | 32 [25, 45] | 13 [12, 14] |  |  |  |
| BMI | 24 [22, 27] | 25 [23, 27] | 21 [18, 24] |  |  |  |
| Insulin modality | Closed loop 226 (47%),<br>Insulin pump 183 (38%),<br>MDI 73 (15%) | Closed loop 160 (44%),<br>Insulin pump 148 (41%),<br>MDI 55 (15%) | Closed loop 66 (56%),<br>Insulin pump 35 (29%), MDI 18 (15%) |  |  |  |
| HbA1c (% and mmol/mol) | 6.8 [6.3, 7.3]<br>51 [45, 56] | 6.7 [6.2, 7.1]<br>50 [44, 54] | 7.1 [6.6, 7.8]<br>54 [49, 62] |  |  |  |
| Years since diagnosis | 12 [9, 20] | 15 [10, 23] | 10 [7, 11] |  |  |  |
| Race | White: 435 (90%), Not Reported: 14 (2.9%), Multiple: 11 (2.3%), Asian: 10 (2.1%), Black/African American: 8 (1.7%), American Indian/Alaskan Native 2 (0.4%), Unknown: 2 (0.4%) | White: 328 (90.4 %), Not reported: 10 (2.8 %), Asian: 9 (2.5 %), Black/African American: 7 (1.9 %), Multiple: 6 (1.7 %), American Indian/Alaskan Native: 2 (0.6 %), Unknown: 1 (0.3 %) | White: 107 (89.9 %), Multiple: 5 (4.2 %), Not reported: 4 (3.4 %), Asian: 1 (0.8 %), Unknown: 1 (0.8 %), Black/African American: 1 (0.8 %) |  |  |  |
| Sex | Female: 329 (68%), Male 153 (32%) | Female 281 (77%), Male 82 (23%) | Male 71 (60%), Female 48 (40%) |  |  |  |
| glycemic Characteristics |  |  |  |  |  |  |
| Average glucose (mg/dL) | 151 [138, 171] | 146 [136, 163] | 170 [150, 187] |  |  |  |
| SD (mg/dL) | 52 [44, 62] | 49 [42, 59] | 58 [48, 68] |  |  |  |
| CV (%) | 34 [30, 37] | 34 [30, 37] | 34 [31, 38] |  |  |  |
| Time in range 70–180 mg/dL (%) | 72 [58, 80] | 73 [62, 82] | 62 [51, 73] |  |  |  |
| Time in range <70 mg/dL (%) | 2.1 [0.8, 3.7] | 2.3 [1.1, 4.1] | 1.0 [0.4, 2.5] |  |  |  |
| Time in range >180 mg/dL (%) | 26 [16, 39] | 23 [15, 34] | 37 [24, 48] |  |  |  |
| Event Characteristics |  |  |  |  |  |  |
| Characteristi | Physical | Non-activity | Physical | Non-activity | Physical | Non-activity |

**Table 1. Participant Characteristics, Baseline glycemic Metrics, and Physical Activity** **Bout Features in the Overall Cohort and by Study Group (T1DEXI vs.**
| **c** | **Activity** |  | **Activity** |  | **Activity** |  |
| --- | --- | --- | --- | --- | --- | --- |
| Number of bouts | 1546 | 1546 | 1293 | 1293 | 253 | 253 |
| Duration of bout (min) | 24 [20, 30] | 24 [20, 30] | 25 [20, 30] | 25 [20, 30] | 20 [15, 30] | 20 [15, 30] |
| Insulin on board (U/kg) | 0.01 [0.00, 0.03] | 0.02 [0.01, 0.03] | 0.01 [0.00, 0.03] | 0.02 [0.00, 0.03] | 0.02 [0.00, 0.04] | 0.02 [0.01, 0.05] |
| Starting glucose (mg/dL) | 210 [193, 235] | 209 [192, 234] | 209 [192, 232] | 208 [192, 232] | 215 [195, 252] | 216 [195, 251] |
| Starting rate of change (mg/dL/ min) | 0.00 [-0.55, 0.70] | 0.04 [-0.50, 0.75] | 0.00 [-0.55, 0.70] | 0.00 [-0.50, 0.70] | 0.04 [-0.59, 0.80] | 0.04 [-0.55, 0.76] |
| CV in the 1 h prior to PA (%) | 5.9 [3.4, 9.6] | 5.7 [3.5, 9.2] | 5.9 [3.4, 9.9] | 5.7 [3.4, 9.4] | 5.6 [3.2, 8.7] | 5.7 [3.7, 8.5] |
| Time of day | Afternoon: 700 (45%),<br>Evening: 456 (30%),<br>Morning: 390 (25%) | Evening: 619 (40%),<br>Afternoon: 467 (30%),<br>Morning: 467 (30%) | Afternoon: 560 (43%),<br>Evening: 398 (31%),<br>Morning: 335 (26%) | Evening: 518 (40%),<br>Afternoon: 402 (31%),<br>Morning: 373 (29%) | Afternoon: 140 (55%),<br>Evening: 58 (23%),<br>Morning: 55 (22%) | Evening: 101 (40%),<br>Morning: 94 (37%),<br>Afternoon: 58 (23%) |
| Type of PA | Aerobic: 875 (57%),<br>Mixed: 537 (35%),<br>Anaerobic: 134 (8%) | N/A | Aerobic: 676 (52%),<br>Mixed: 494 (39%),<br>Anaerobic: 123 (9%) | N/A | Aerobic: 199 (79%),<br>Mixed: 43 (17%),<br>Anaerobic: 11 (4%) | N/A |
| Intensity | Light: 768 (50%),<br>Moderate: 653 (42%),<br>Vigorous: 125 (8%) | N/A | Light: 579 (45%),<br>Moderate: 602 (47%),<br>Vigorous: 112 (8%) | N/A | Light: 189 (75%),<br>Moderate: 51 (20%),<br>Vigorous: 13 (5%) | N/A |

### Matching and PA Bout Inclusion

A comprehensive set of individual and event-level variables was assessed for their influence on glucose responses to PA. Individual-level variables included insulin modality, BMI, and HbA1c. Event-level variables included PA type, intensity, time of day, insulin dose, glucose rate of change, insulin on board (IOB), activity duration, starting glucose level, and glucose coefficient of variation (CV) in the hour before activity. Following hierarchical modelling, the final matching algorithm retained four key variables: starting glucose, glucose rate of change, IOB, and pre-activity glucose CV. Of the 1,944 eligible PA bouts, 398 were excluded due to insufficient match quality based on predefined thresholds. The remaining 1,546 matched pairs were evaluated using SMDs, with SMDs <0.1 indicating successful covariate balance across matched variables (20).

### Characteristics at the Start of Physical Activity Bouts

Of the 1,546 bouts, 1,293 were from the T1DEXI cohort, while 253 were from the T1DEXIP cohort. Full pre-PA characteristics are available in **Table 1**.

The overall median duration of PA bouts was 23 minutes (IQR: 20, 30), with a median of 25 minutes (20, 30) for T1DEXI participants and 20 minutes (15, 30) for those in the T1DEXIP group. Starting glucose level before activity for the total cohort was 210 mg/dL [193, 235] (11.7 mmol/L [10.7, 13.1]), with it being 209 mg/dL [192, 232] (11.6 mmol/L [10.7, 12.9]) for the T1DEXIgroup and 215 mg/dL [195, 252] (11.9 mmol/L [10.8, 14.0]) for the T1DEXIP. Median IOB was comparable across groups, with T1DEXI having 0.01 U/kg (0.0, 0.03) and the T1DEXIP having 0.02 U/kg (0.0, 0.04) at PA onset.

In the T1DEXI cohort, 45% of PA bouts were light, 47% moderate, and 8% vigorous intensity. For the T1DEXIP group, 75% were light, 20% moderate, and 5% vigorous intensity. For PA modality, the T1DEXI performed a mix of aerobic (52%), mixed (39%), and anaerobic (9%) activity types. The T1DEXIP engaged in aerobic (79%), mixed (17%), and anaerobic (4%) activities. T1DEXI bouts occurred most frequently in the afternoon (43%), followed by the evening (31%), and then the morning (26%). For T1DEXIP, the afternoon was also most common (55%), with fewer bouts in the evening (23%) and morning (22%).

### Primary Outcome

#### Effect of PA on Glucose Change

In the mixed-effects model, after adjusting for relevant covariates and accounting for clustering by participant and matched bout, PA was associated with a significantly greater reduction in glucose compared with matched non-PA periods (mean difference −35 mg/dL [95% CI −37 to −32]; −1.9 mmol/L [−2.1 to −1.8]; p < 0.001). The mean glucose reduction following PA was −40 mg/dL (95% CI −43 to −35; −2.2 mmol/L [−2.4 to −1.9]; p < 0.001), compared with −5 mg/dL (95% CI −7 to −3; −0.3 mmol/L [−0.4 to −0.2]; p < 0.001) following matched non-PA periods. The glucose response for each PA bout, ordered by magnitude of drop and compared with its matched non-activity period, is available in the **Supplementary Data (S4).**

The model included random intercepts for both participants and matched PA/non-PA periods, allowing each participant and each period to have its baseline level of the outcome. The variance attributed to participants (0.37±0.61) reflects baseline differences in glucose change across individuals, while the smaller variance associated with matched bouts (0.01±0.10) suggests minimal unexplained variability between paired observations. The residual variance remained substantial (4.23±2.06), consistent with known intra-individual and physiological variability in glucose responses. Together, these random effects appropriately accounted for clustering by participant and the matched design, supporting the use of a multilevel structure in the model.

#### Predictors of the differential glucose-lowering effect of PA

Further analyses examined factors modifying the effect of PA on glucose reduction. The glucose-lowering effect of PA was greater in bouts with a faster rate of glucose decline at onset (per 1.8 mg/dL/min: −16.2 mg/dL [95% CI −21.4 to −11.0]; −0.90 mmol/L [−1.19 to −0.61]; p < 0.001) (**Fig. 2A**), lower starting glucose (per 18 mg/dL: −2.9 mg/dL [95% CI −4.1 to −1.6]; −0.16 mmol/L [−0.23 to −0.09]; p < 0.001) (**Fig. 2B**), higher pre-PA CV (per 1%: −1.1 mg/dL [95% CI −1.6 to −0.5]; −0.06 mmol/L [−0.09 to −0.03]; p < 0.001) (**Fig. 2C**), longer duration (per minute: −0.9 mg/dL [95% CI −1.3 to −0.5]; −0.05 mmol/L [−0.07 to −0.03]; p < 0.001) (**Fig. 2D**), and higher IOB at onset (per 0.1 U/kg: −10.1 mg/dL [95% CI −19.6 to −0.7]; −0.56 mmol/L [−1.09 to −0.04]; p = 0.035) (**Fig. 2E**).

**Figure 2.**
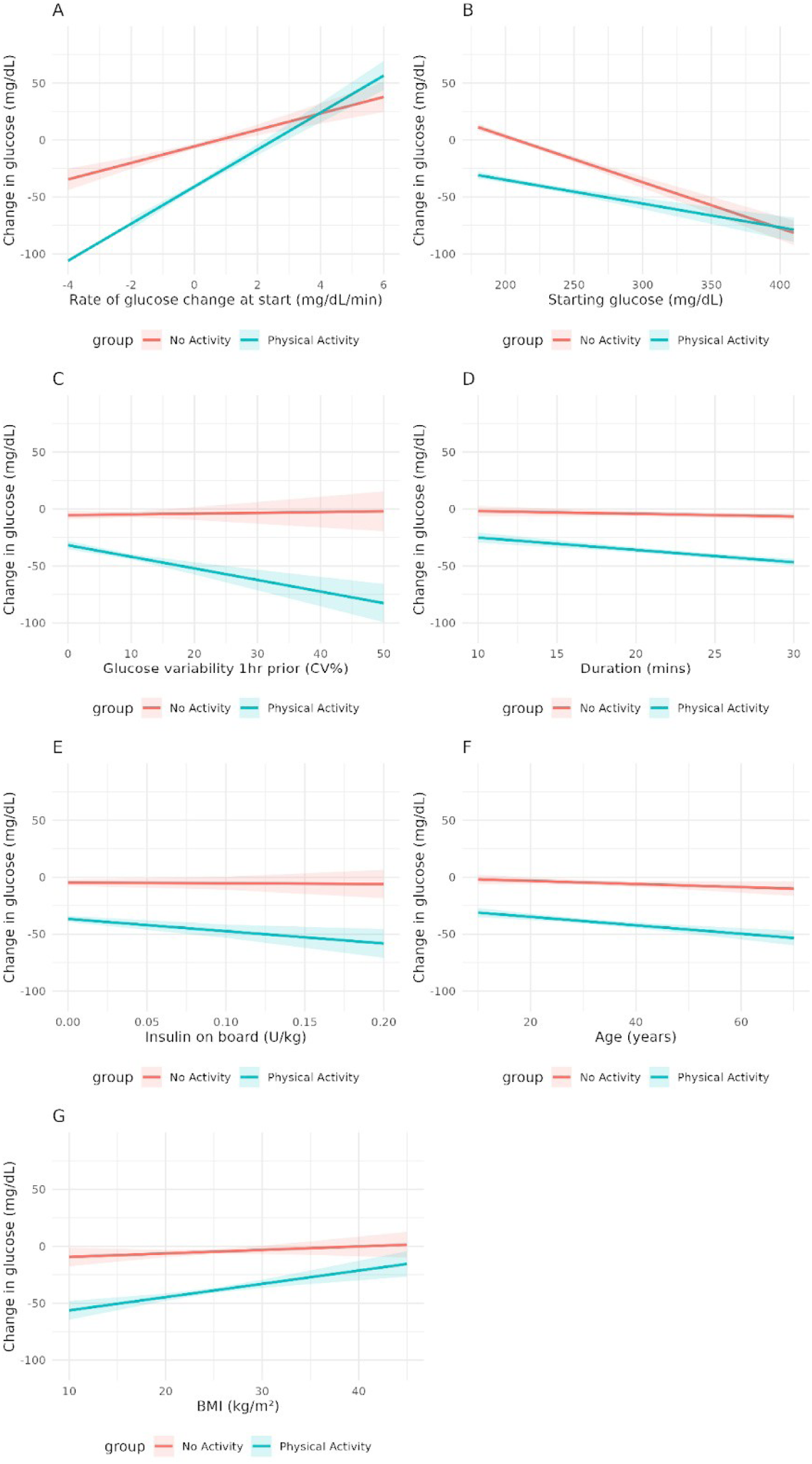
Individual- and Event-Level Moderators of the Differential Glucose Drop Between Physical Activity and Non-PA. This figure displays individual- and event-level factors that influenced the difference in glucose change between physical activity (PA) and matched non-PA periods: (A) starting glucose rate of change (B) starting glucose level (C) Pre-PA (1-hour) glucose coefficient of variation (CV) (D) Activity duration (E) starting insulin on board (IOB) (F) Age (G) Body mass index (BMI).

Older age (per year: −0.2 mg/dL [95% CI −0.4 to 0.0]; −0.01 mmol/L [−0.02 to 0.00]; p = 0.015) (**Fig. 2F**) and lower BMI (per 1 kg/m²: −0.9 mg/dL [95% CI −1.6 to −0.2]; −0.05 mmol/L [−0.09 to −0.01]; p = 0.012) were also associated with greater reductions. The model included random intercepts for participants and matched bouts. Participant-level variance (0.37±0.61) reflected baseline differences in glucose change, while variance for matched bouts was minimal (0.01±0.10). Residual variance was 4.23±2.06, consistent with intra-individual variability. The model output is available in the **Supplementary Data (S3)**.

### Heatmap Decision Tool

The three strongest predictors of glucose change, PA status (defined as whether 10–30 minutes of activity was undertaken), starting glucose level, and rate of glucose change at event onset, were used to model predicted glucose patterns (**Fig. 3**). This visual tool shows how glucose responses vary depending on whether or not PA is performed.

**Figure 3.**
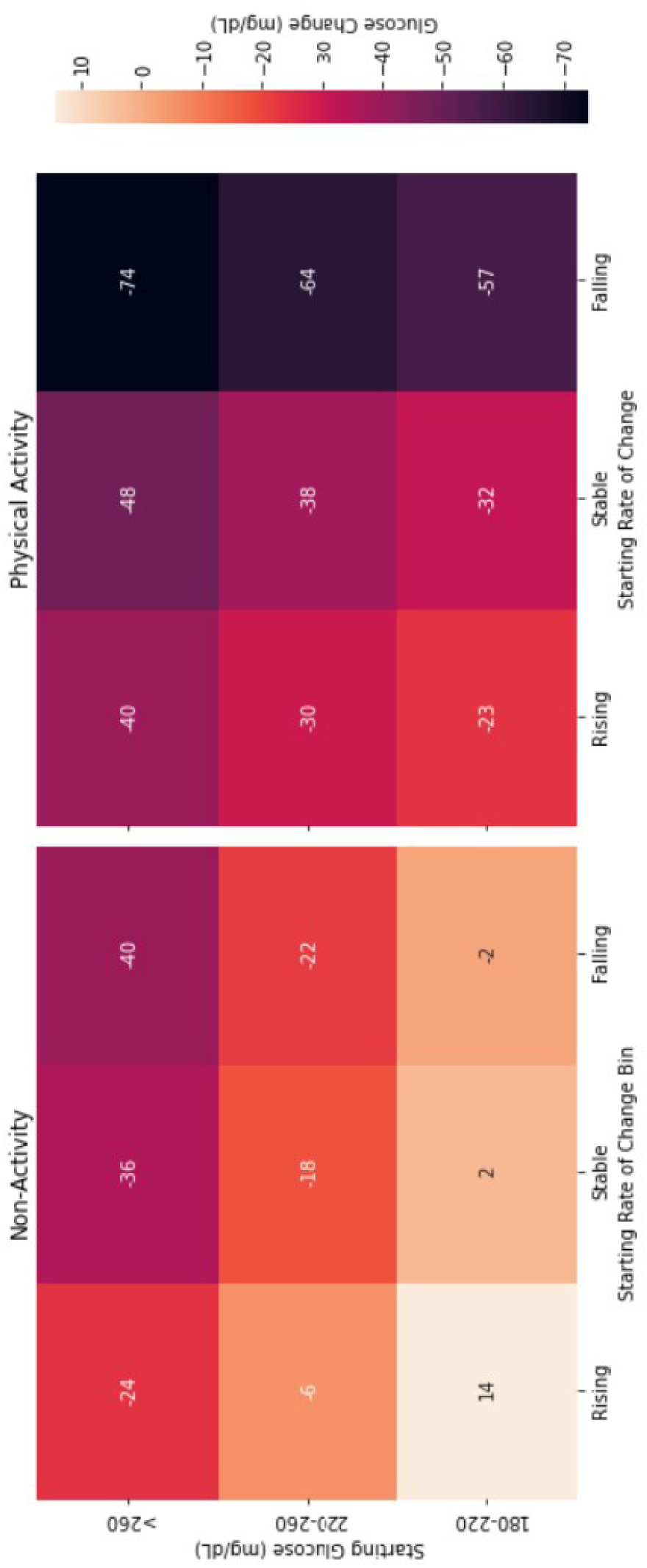
Predicted Glucose Trajectories Based on Physical Activity Status, Starting Glucose, and Glucose Rate of Change. This heatmap illustrates predicted glucose changes following a 23-minute (median) event with 20-minutes post-event based on three key variables: physical activity status, starting glucose level, and starting glucose rate of change. The figure shows how glucose responses differ when physical activity (PA) is undertaken versus not, across a range of starting glucose levels and trends. Warmer colors indicate greater predicted glucose reductions during PA.

For example, with a starting glucose of 180–220 mg/dL (10.0–12.2 mmol/L) and a stable (→) trend arrow (glucose rate of change between −1 and 1 mg/dL/min; −0.06 to 0.06 mmol/L/min), glucose is expected to rise by 2 mg/dL (0.1 mmol/L) over 20 minutes without PA, but to fall by 38 mg/dL (2.1 mmol/L) with PA, a 40 mg/dL (2.2 mmol/L) difference. At higher starting levels (220–260 mg/dL; 12.2–14.4 mmol/L) and a falling (↘ or ↓) trend (glucose rate of change < −1 mg/dL/min; < −0.06 mmol/L/min), the predicted drop is 22 mg/dL (1.2 mmol/L) without PA and 64 mg/dL (3.6 mmol/L) with PA, a 42 mg/dL (2.3 mmol/L) difference.

### Secondary Outcomes: Hypoglycemia Risk

The absolute risk of hypoglycemia during and up to 20 minutes after was 1.4% for PA bouts (22 out of 1,546) and 0.1% for matched non-PA periods (1 out of 1,546). All of the hypoglycemic episodes occurred in the adult T1DEXI dataset. Although the overall risk was low, PA was strongly associated with a higher likelihood of hypoglycemia. In the fully adjusted generalised linear mixed model (binomial), PA was associated with a log-odds increase of 4.48, corresponding to an odds ratio (OR) of 88 (95% CI: 88, 89, p < 0.001). Two additional factors were significantly associated with hypoglycemia risk: starting glucose (log-odds = −0.57, OR = 0.56) and event duration (log-odds = 0.15, OR = 1.16). Other variables, including age, BMI, rate of glucose change at event onset, and insulin delivery modality, were not significantly associated with risk. Full model results are provided in the **Supplementary Data (S5).**

### Sensitivity Analysis

To test the reliability of our results, we conducted a sensitivity analysis by removing all bouts where participants recorded eating carbohydrates and/or taking insulin during the measurement period (n = 303). This was done to isolate the effect of PA on glucose levels without interference from external glucose or insulin.

When these bouts were excluded, the average glucose drop during PA was slightly smaller (−36 mg/dL; 2.0 mmol/L), and no hypoglycemia bouts were observed (0 of 1,243). This suggests that participants who consumed carbohydrates were likely reacting to a rapid glucose drop, and excluding these bouts could underestimate the true effectiveness and safety of PA for lowering glucose.

## Conclusions

This study provides strong evidence that 10–30 minute bouts of PA significantly accelerate glucose lowering in people with T1D compared to matched periods without PA, when individuals would typically rely on insulin correction or passive glucose decline. Using a within-subject matched-pairs design and 1,546 free-living PA bouts, we found that a median of 23 minutes of PA, initiated when glucose was above 180 mg/dL, lowered glucose by an average of 40 mg/dL (2.2 mmol/L), compared to just 5 mg/dL (0.3 mmol/L) during matched non-PA periods. This represents an eightfold greater reduction attributable to PA, consistent across a diverse cohort of adolescents and adults using various insulin delivery methods and engaging in different types and intensities of activity. These findings demonstrate that even relatively brief periods of PA, using any form or intensity, can serve as an effective and practical strategy to rapidly lower elevated glucose levels.

Causal inference methods are increasingly recognized as essential in diabetes research, particularly for generating real-world evidence where even crossover study designs may not fully address confounding or reflect everyday conditions. Previous studies have shown that carefully matched observational data can mimic trial conditions to estimate treatment effects (26). For example, Deng et al. (27) used this approach to closely replicate the GRADE trial, comparing glucose-lowering therapies in people with type 2 diabetes using real-world data. Our study builds on this framework by applying causal methods to PA and glucose control in people with T1D. Using a within-subject matched-pairs design, each PA event was paired with a matched non-PA period from the same individual. This allowed us to control for fixed factors like age, sex, insulin regimen, and usual activity patterns, while also balancing key variables that change over time, such as starting glucose, glucose rate of change, CV, and IOB.

This design closely mimics a ‘what-if?’ comparison and strengthens causal inference by increasing confidence that the glucose-lowering effects were due to PA itself, not other behavioural or metabolic factors. This approach marks a significant improvement over previous research using the T1DEXI and T1DEXIP datasets, which showed associations between PA and improved glucose but could not robustly infer causality (11).

Our findings provide the strongest evidence to date that bouts of PA are a very safe, fast-acting, and effective way to lower high glucose levels. The effect was consistent across different times of day, age groups, and insulin delivery methods, underscoring its broad clinical relevance and supporting its potential as a foundation for future interventions and clinical guidance. Notably, the risk of hypoglycemia was low. These results support the use of short PA sessions to manage hyperglycemia and may help reduce the fear of hypoglycemia, which is commonly reported by people living with T1D (28,29).

We identified starting glucose and glucose rate of change as the strongest predictors of glucose lowering with PA. A steeper downward glucose rate of change at activity onset led to a greater reduction, while a rising trend reduced the effect. Together, these variables explained much of the variation in glucose outcomes during both PA and matched non-PA periods. This aligns with previous findings from the T1DEXI and T1DEXIP studies, where these factors were key predictors of glycemic response to activity (16,30). Our study builds on this by confirming the associations using causal modelling and introducing a heatmap tool for clinical use. This visual tool shows how starting glucose and rate of change, readily available from CGM, can guide personalised, real-time decisions, refining the general finding that ∼20 minutes of PA can lower glucose by ∼40 mg/dL (2.0 mmol/L).

IOB was a key factor influencing the glucose-lowering effect of PA. Higher IOB at activity onset significantly enhanced glucose reductions, consistent with physiological mechanisms whereby PA increases insulin sensitivity, accelerates subcutaneous insulin absorption, and reduces renal insulin clearance (8,9). This aligns with a systematic review showing that postprandial PA lowers glucose by 36–144 mg/dL (2–8 mmol/L), depending on timing and intensity (10). Controlled trials also show that even with 25– 75% reduced insulin dosing in the 2–4 hours prior, PA can restore glucose to below 180 mg/dL (10 mmol/L) within 30–60 minutes (31–33). Despite its clinical and research relevance, IOB remains challenging to quantify. In this study, it was estimated using a 4- hour linear decay model, which underestimates insulin action. Insulin can continue to act for five to six hours (34), yet real-world bolus calculators often assume shorter durations and overlook individual differences, limiting their utility for PA decisions (5).

The absence of a significant effect of activity type or intensity is both surprising and encouraging. It suggests that individuals with T1D can engage in a wide variety of activities and still achieve clinically meaningful glucose reductions. This flexibility may enhance adherence and allow for more personalized, lifestyle-integrated approaches to glucose management through PA.

Personal factors also influenced the glucose-lowering effect of PA. Participants with lower BMI had greater glucose reductions, likely due to higher baseline insulin sensitivity. This is consistent with previous findings from the T1DEXI and T1DEXIP cohorts (16,30). Those with higher HbA1c levels showed smaller reductions, suggesting that chronic hyperglycemia and insulin resistance may blunt the effectiveness of PA. Higher baseline fitness also appeared to enhance glucose response, which is supported by mechanistic studies showing that regular PA improves insulin action by increasing GLUT4 expression and reducing insulin resistance (8). Age was a modest moderator, with older participants experiencing slightly greater glucose drops. One possible explanation is that younger individuals may consume carbohydrates before activity due to fear of hypoglycemia, reducing the observed effect. However, this remains speculative and requires further study.

This study has several important strengths. It is the first large-scale analysis to use a within-subject matched-pairs causal design to evaluate the acute effect of PA on hyperglycemia in people with T1D using real-world CGM data. Matching each PA bout to a non-PA bout from the same individual controlled for fixed person-level and key time-varying factors, strengthening causal inference and providing a robust estimate of PA’s independent effect on postprandial glycemia. The analysis included over 1,500 matched pairs across more than 400 participants, spanning a wide age range, insulin delivery methods, and types and intensities of PA. Harmonized data from two well-characterized cohorts enabled consistent processing and integration of sensor, insulin, and self-reported activity information. A sensitivity analysis confirmed that retaining the largest dataset was crucial to avoid introducing unintended bias.

Several limitations should be acknowledged. First, although matching reduces bias, residual confounding may remain, particularly from unmeasured factors such as sleep, stress, or unrecorded food intake. Second, PA was self-reported in the T1DEXIP cohort and device-measured in the T1DEXI cohort, potentially introducing variability in event classification or intensity estimates. Third, IOB was estimated using a simplified 4-hour linear decay model that does not fully reflect rapid-acting insulin pharmacodynamics.

Fourth, the study focused on 10–30-minute PA and glucose effects up to 20 minutes post-activity, limiting assessment of longer-term outcomes like two-hour AUC and delayed hypoglycemia. Lastly, given the homogeneous cohort and varied PA modes and intensities, further research is needed across more diverse populations.In summary, this study provides strong evidence that short bouts of PA are a very safe and effective way to lower high glucose levels in people with type 1 diabetes. A 20- minute session lowered glucose by an average of ∼40 mg/dL (2 mmol/L), which could be clinically translated into a simple “40-every-20” or “2-every-20” rule. The strong influence of starting glucose and glucose rate of change supports the use of heatmaps and digital tools to personalize this strategy in education and clinical care. Future research should explore whether integrating this approach into care pathways and digital platforms can improve time in range, reduce insulin use, and support safer physical activity.

## Supporting information

STROBE Guidelines

## Data Availability

The Helmsley datasets (T1-DEXI and T1-DEXIP) used in this study are available via application through Vivli (https://vivli.org), a global clinical research data sharing platform.

## Author Contributions

Catherine L. Russon: Background research, Formal analysis, Writing – original draft. John Pemberton: Conceptualization, Background research, Writing – original draft.

Richard M. Pulsford: Analysis, Writing – review & editing. Bradley S. Metcalf: Analysis, Writing – review & editing. Emma Cockroft: Writing – review & editing.

Michael Allen: Analysis, Writing – review & editing. Anne-Marie Frohock: Writing – review & editing.

Robert C. Andrews: Supervision, writing – review & editing, intellectual revision.

## Data Access and Responsibility: Guarantor Statement

Rob C. Andrews, Catherine L. Russon and John Pemberton are the guarantors of this work and, as such, had full access to all the data in the study and take responsibility for the integrity of the data and the accuracy of the data analysis.

## Data Availability Statement

The data that support the findings of this study are available from the corresponding author upon reasonable request.

## Funding and Assistance

### Original Study Support

This study was conducted as part of the T1DEXI Initiative, supported by the Leona M. and Harry B. Helmsley Charitable Trust.

### Non-Financial Contributions

Verily Life Sciences (South San Francisco, CA) provided the Study Watch devices at no cost. Dexcom Inc. supplied continuous glucose monitoring systems at a discounted rate.

## Acknowledgments

We would like to sincerely acknowledge the invaluable contributions of the following collaborators for their ongoing discussion, insight, and foundational work in developing the concept of using physical activity to lower glucose in individuals with type 1 diabetes. In particular, we wish to highlight their pioneering efforts in curating and analysing the T1DEXI and T1DEXIP datasets, which have provided the critical infrastructure to explore this question through real-world, high-resolution data: Zoey Li (Jaeb Center for Health Research, Tampa, FL, USA), Robin L. Gal (Jaeb Center for Health Research, Tampa, FL, USA), Lauren V. Turner (School of Kinesiology and Health Science, York University, Toronto, ON, Canada), Simon Bergford (Jaeb Center for Health Research, Tampa, FL, USA), Peter Calhoun (Jaeb Center for Health Research, Tampa, FL, USA), and Michael C. Riddell (School of Kinesiology and Health Science, York University, Toronto, ON, Canada). Their sustained support has been instrumental in shaping the research direction and ensuring its translational relevance.

## AI Use Disclosure

During the course of preparing this work, the authors used OpenAI DALL·E 3, accessed via ChatGPT platform, OpenAI, San Francisco, CA; April 2025) for the purpose of graphical abstract creation and to refine grammar, improve readability, and enhance clarity in the manuscript text.. Following the use of this tool/service, the authors formally reviewed the content for its accuracy and edited it as necessary. The authors take full responsibility for all the content of this publication.

## Prior Presentation

Preliminary analyses from this study were presented at the Diabetes UK Professional Conference, Glasgow, UK, in February 2025. A preprint version of this manuscript was also uploaded to medRxiv on May 2nd, 2025

