## Supplementary Data for "Glucose-lowering effects of physical activity in type 1 diabetes: A causal modelling and matched-pair analysis approach"

**S1. Balance Diagnostics for Matched Physical Activity and Non-Activity Bouts.** *Standardised mean differences (SMDs) were used to assess balance between matched physical activity (PA) and non-PA bouts across key variables. An SMD <0.1 was considered indicative of good balance. Variables include demographic characteristics, clinical features, and bout-specific glucose and insulin measures. Balance diagnostics were conducted after matching to ensure comparability of activity and non-activity events.*

| **Variable** | **SMD** |
| --- | --- |
| IOB/kg | 0.090 |
| CV | 0.023 |
| Start glucose | 0.017 |
| Start rate of change | 0.007 |

**S2. Linear Mixed-Effects Model Predicting Change in Glucose Following Physical Activity Events.** *Results are from a linear mixed-effects model assessing the change in glucose (mg/dL) following physical activity (PA) bouts, with random intercepts for bout ID and participant ID. Fixed effects include demographic variables, bout characteristics, and their interactions with PA. All predictors were mean-centred but not standardised to preserve interpretability in original units. Estimated coefficients are reported with 95% confidence intervals*

| **Term** | **Estimate [95 % CI]** | **p-value** |
| --- | --- | --- |
| (Intercept) | −0.27 [−0.39, −0.15] | 1.34 × 10⁻⁵ |
| Exercise | −1.91 [−2.05, −1.77] | < 2 × 10⁻¹⁶ |
| Duration | −0.013 [−0.030, 0.004] | 0.127 |
| Start ROC | 7.25 [5.02, 9.47] | 1.95 × 10⁻¹⁰ |
| Age | −0.0076 [−0.016, 0.0006] | 0.089 |
| CV | 0.0038 [−0.018, 0.026] | 0.744 |
| BMI | 0.017 [−0.013, 0.048] | 0.265 |
| IOB (U kg⁻¹) | −0.34 [−4.25, 3.57] | 0.864 |
| Starting glucose | −0.40 [−0.46, −0.35] | < 2 × 10⁻¹⁶ |
| HbA1c | 0.031 [0.020, 0.042] | 5.97 × 10⁻⁸ |
| Exercise × Duration | −0.047 [−0.069, −0.025] | 4.07 × 10⁻⁵ |
| Exercise × Start ROC | 9.02 [6.02, 12.02] | 4.35 × 10⁻⁹ |
| Exercise × Age | −0.013 [−0.024, −0.002] | 0.015 |
| Exercise × CV | −0.060 [−0.091, −0.029] | 9.69 × 10⁻⁵ |
| Exercise × BMI | 0.048 [0.011, 0.085] | 0.0115 |
| Exercise × IOB (U kg⁻¹) | −5.64 [−10.89, −0.39] | 0.035 |
| Exercise × Glucose | 0.195 [0.123, 0.267] | 1.36 × 10⁻⁷ |

**S3. Random Effects from the Linear Mixed-Effects Model Predicting Glucose Change Following Physical Activity.** *This table presents the variance components and standard deviations for the random effects included in the mixed-effects model described in Table 3. Random intercepts were specified for both physical activity bout ID and participant ID to account for repeated measures within individuals and within activity events. The residual variance reflects unexplained variability after accounting for fixed and random effects.*

| **Group** | **Effect** | **Variance** | **Std. Dev.** |
| --- | --- | --- | --- |
| bout_id | Intercept | 3.469 | 1.863 |
| ID | Intercept | 119.067 | 10.912 |
| Residual | — | 1371.865 | 37.039 |

**S4. Mean ± SD Glucose Change per Participant (≥3 PA Events): Exercise vs Matched Non-Exercise Bouts*.*** *This figure displays the mean change in glucose (mg/dL) for participants with ≥3 physical activity (PA) bouts, comparing PA (red) to matched non-PA bouts (blue). Each point represents a participant’s average glucose change, with horizontal lines indicating ±1 standard deviation. Participants are ordered by the magnitude of their average glucose drop during exercise. This plot illustrates considerable inter-individual variability, with most participants experiencing a greater glucose-lowering effect during PA than in matched non-activity periods.*

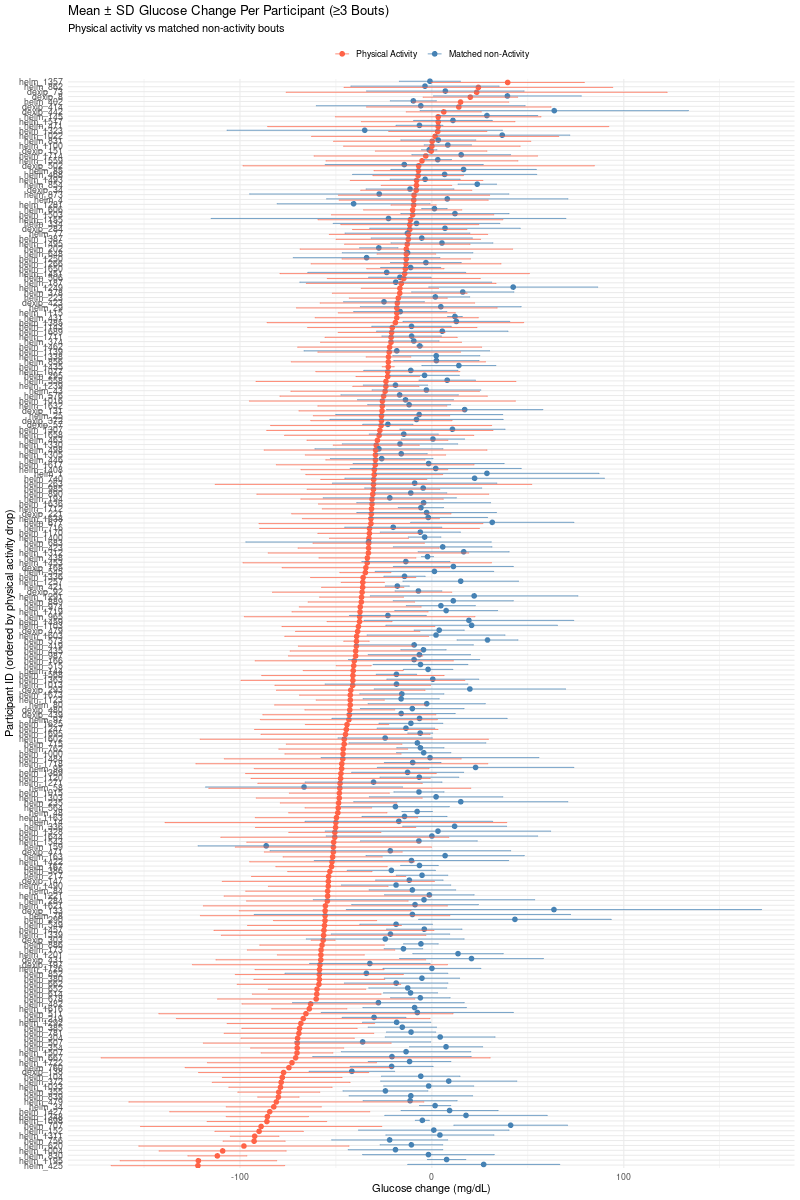

**S5. Summary of Variables Included in the Hypoglycemia Risk Model.** *This table provides a detailed overview of variables used in the hypoglycemia analysis pipeline. Each variable is listed alongside its type, unit (where applicable), and a brief description. Variables include participant demographics, bout characteristics, glycaemic metrics, and derived predictors (e.g. glucose rate of change, insulin on board). Variables were derived from CGM and pump data collected around physical activity events.*

| **Variable** | **Brief description** |
| --- | --- |
| form_of_exercise | Categorical label for whether the physical activity was aerobic, anaerobic or mixed |
| time_of_day | When the session began—typically coded as morning / afternoon / evening or a clock time |
| day_of_week | Day on which the session occurred (Monday … Sunday) |
| insulin_modality | Therapy type used for the session (e.g., MDI vs pump) |
| sex | Participant’s biological sex |
| food_pre_30 | Grams of carbohydrate eaten ≤ 30 min before exercise |
| food_pre_60 | Grams of carbohydrate eaten 30–60 min before exercise |
| iob_kg | Insulin on board per kilogram at the start of physical activity |
| ins_hrs | Number of hours since last insulin bolus |
| start_glc | Starting glucose at exercise onset (mmol L⁻¹) |
| start_roc | Glucose rate-of-change at onset (mmol L⁻¹ min⁻¹) |
| time_since_last_ins_dose | Minutes since last insulin bolus/basal adjustment |
| duration | Exercise bout length (minutes) |
| intensity | Intensity measure (e.g., %HRmax, METs, RPE score) |
| hba1c | Glycated haemoglobin—long-term glycaemic control (mmol/mol) |
| bmi | Body-mass index (kg m⁻²) |
| age | Participant age (years) |
| time_since_last_activity | Hours since the previous structured physical activity |
| Average glucose (mmol/L) | Mean glucose over the analysis window in the 1hr prior to exercise |
| SD (mmol/L) | Standard deviation of glucose values in the 1hr prior to exercise |
| CV (%) | Coefficient of variation of glucose (SD ÷ mean × 100) in the 1hr prior to exercise |
| AUC (mmol h L⁻¹) | Area under the glucose-time curve in the 1hr prior to exercise |
| LBGI | Low Blood Glucose Index—aggregate hypoglycaemia risk in the 1hr prior to exercise |
| HBGI | High Blood Glucose Index—aggregate hyperglycaemia risk in the 1hr prior to exercise |
| MAGE (mmol/L) | Mean Amplitude of Glycaemic Excursions in the 1hr prior to exercise |
| TIR normal (%) | % of time 3.9–10 mmol L⁻¹ (primary “time-in-range”) in the 1hr prior to exercise |
| TIR level 1 hypoglycemia (%) | % time 3.0–3.8 mmol L⁻¹ (“level 1” hypo) in the 1hr prior to exercise |
| TIR level 2 hypoglycemia (%) | % time < 3.0 mmol L⁻¹ (“level 2” hypo) in the 1hr prior to exercise |
| TIR level 1 hyperglycemia (%) | % time 10.0–13.9 mmol L⁻¹ (“level 1” hyper) in the 1hr prior to exercise |
| TIR level 2 hyperglycemia (%) | % time ≥ 14.0 mmol L⁻¹ (“level 2” hyper) in the 1hr prior to exercise |
| Total number hypoglycemic events | Count of all hypoglycaemic episodes in the 1hr prior to exercise |
| Number LV1 hypoglycemic events | Count of level 1 hypoglycaemic episodes in the 1hr prior to exercise |
| Number LV2 hypoglycemic events | Count of level 2 hypoglycaemic episodes in the 1hr prior to exercise |
| Number prolonged hypoglycemic events | Hypo events exceeding the set length threshold in the 1hr prior to exercise |
| Total number hyperglycemic events | Count of all hyperglycaemic episodes in the 1hr prior to exercise |
| Number LV1 hyperglycemic events | Count of level 1 hyperglycaemic episodes in the 1hr prior to exercise |
| Number LV2 hyperglycemic events | Count of level 2 hyperglycaemic episodes in the 1hr prior to exercise |
| Number prolonged hyperglycemic events | Hyper events exceeding the set length threshold in the 1hr prior to exercise |
