## Supplementary material for "Glucose-lowering effects of physical activity in type 1 diabetes: A causal modelling and matched-pair analysis approach": STROBE Guidelines

STROBE Checklist with Descriptions and Page Numbers

| STROBE Item | Description of How Item Was Met | Page(s) in Manuscript |
| --- | --- | --- |
| Title and Abstract | (a) The abstract clearly identifies this as a "within-subject matched-pairs causal design" study. (b) The abstract provides a balanced summary of the study’s aims, methods, results, and implications. | Abstract (p.1) |
| Background/Rationale | The introduction explains the burden of hyperglycemia in T1D and the limitations of insulin correction, establishing the need for alternative approaches such as physical activity. | Introduction (p.2) |
| Objectives | The study explicitly aims to evaluate the glucose-lowering effects of bouts of PA using a causal inference framework and matched control design. | Introduction (p.2) |
| Study Design | A within-subject matched-pairs observational study was used, applying causal inference techniques to CGM data. | Research Design and Methods (p.3) |
| Setting | The setting involved free-living environments using data from T1DEXI and T1DEXIP cohorts. Data were collected via CGM and accelerometer devices. No specific recruitment window is required due to the retrospective nature. | Research Design and Methods (p.3) |
| Participants | (a) Eligibility criteria: use of CGM, insulin, and accelerometer data with >70% completeness; PA initiated at glucose >180 mg/dL, n=482 (n=363 adults, n=119 adolescents) (b) Matching criteria: starting glucose, glucose rate of change, IOB, CV; n=1546, matched pairs. | Research Design and Methods (p.3-4) |
| Variables | Outcomes: change in glucose post-PA; exposures: PA vs. matched non-PA periods; confounders and modifiers: IOB, starting glucose, glucose rate of change, BMI, etc. | Research Design and Methods (p.4) |
| Data Sources/Measurement | T1DEXI and T1DEXIP cohorts; sensor glucose, insulin, and accelerometer data. Standardised methods across participants. | Research Design and Methods (p.4) |
| Bias | Within-subject design, outcome-oriented matching, and use of robust causal matching minimise confounding and selection bias. | Research Design and Methods (p.5) |
| Study Size | Initial PA bouts (n=1,943) were reduced to 1,546 matched pairs based on match quality criteria. | Research Design and Methods (p.5) |
| Quantitative Variables | All continuous variables (e.g., glucose, glucose rate of change, IOB) included in regression models; no arbitrary grouping. | Statistical Analysis (p.5-6) |
| Statistical Methods | (a) Linear/logistic mixed-effects models used to compare glucose change and hypoglycemia odds. (b) Subgroup analyses included moderators (age, BMI, etc.). (c) Missing data addressed through exclusion of poorly matched pairs. (d) Not applicable—study used observational datasets with complete follow-up periods. (e) Matching thresholds and variable weighting were sensitivity-tuned. | Statistical Analysis (p.6) |
| Participants (Results) | (a) 531 participants included, yielding 1,944 PA bouts. (b) 107 PA bouts excluded due to insufficient matching. (c) A flow diagram provided to show inclusions/exclusions | Results (p.7) |
| Descriptive Data | (a) Participant demographics, insulin modality, and glycemic metrics detailed in Table 1. (b) No missing data reported for key variables. (c) Follow-up time not relevant due to short-term outcome. | Table 1 & Results (p.8) |
| Outcome Data | Glucose changes and hypoglycemia outcomes are reported with means and confidence intervals. | Results (p.8-9) |
| Main Results | (a) Estimates presented with 95% CIs; adjusted for key covariates. (b) Continuous variables were retained as-is. (c) Absolute hypoglycemia risk (1.4%) reported alongside OR. | Results (p.9) |
| Other Analyses | Subgroup and moderator analyses performed and visualized using heatmaps and stratified effects. | Results (p.10) |
| Key Results | The glucose-lowering effect of PA was significant and robust across subgroups, summarized clearly in the discussion. | Discussion (p.11) |
| Limitations | Limitations include potential residual confounding, simplified IOB estimates, and absence of dietary data. | Discussion (p.12) |
| Interpretation | Results are cautiously interpreted, contextualized with prior literature, and practical implications are highlighted. | Discussion (p.12-13) |
| Generalisability | Real-world design, broad age range, and varied insulin modalities support generalisability. | Discussion (p.13) |
| Funding | Study supported by the Helmsley Charitable Trust. Devices provided at low/no cost by Verily and Dexcom. Funders had no role in analysis or reporting. | Acknowledgements (p.14) |
